# Depressive Symptom Trajectories and Incident Cognitive Impairment by Outcome Ascertainment

**DOI:** 10.64898/2026.11.18.26368449

**Authors:** Xiaozheng Liu, Zhongwei Guo

## Abstract

**Importance:** Dementia prevention frameworks treat depression as a leading modifiable risk factor, but cohort estimates of the association vary widely; whether this variation reflects how cognitive outcomes are ascertained is unknown.

**Objective:** To examine whether associations between depressive symptom trajectories and incident cognitive impairment vary by outcome ascertainment rigor.

**Design, Setting, and Participants:** This harmonized cohort study included 91,043 adults aged 50 years or older from 4 national longitudinal aging cohorts (CHARLS [China], HRS [US], ELSA [England], and SHARE [16 European countries]); baseline waves spanned 2006 to 2011, with follow-up through 2023.

**Exposure:** Depressive symptom trajectory classes (persistent low, persistent high, incident worsening, remitting) derived using group-based trajectory modeling.

**Main Outcomes and Measures:** Incident cognitive impairment ascertained under a 5-level hierarchy of increasing rigor, from self-report (level 1) to clinical consensus diagnosis (level 5; Harmonized Cognitive Assessment Protocol [HCAP]). Cohort-specific Cox models were pooled by random-effects meta-analysis; meta-regression tested ascertainment rigor as a moderator of heterogeneity.

**Results:** Under nonadjudicated outcomes, the incident-worsening trajectory was associated with incident cognitive impairment in CHARLS (HR, 1.31; 95% CI, 1.06-1.62) and SHARE (HR, 1.99; 95% CI, 1.87-2.13) but not HRS (HR, 1.05; 95% CI, 0.95-1.16) or ELSA (HR, 1.10; 95% CI, 0.95-1.27); the persistent-high trajectory was not significant under any nonadjudicated outcome (HR range, 1.07-1.15). Under clinical consensus diagnosis (n = 3,196), all non-low trajectories were independently associated with mild cognitive impairment or dementia (OR range, 1.66-2.29; dementia-only persistent high: OR, 3.32; 95% CI, 1.85-5.95). Effect sizes increased stepwise with ascertainment rigor (meta-regression slope, 0.314 log HR per level; P < .001; residual I^2^ = 0%; R^2^ = 100%; exploratory). Findings were robust to lagged, weighted, and competing-risk analyses; biological aging acceleration did not differ across trajectories.

**Conclusions and Relevance:** The detectability and magnitude of depression-related cognitive risk depend on outcome ascertainment rigor. Prevention trials and burden projections relying on self-reported outcomes may underestimate the benefit of depression treatment.

**Key Points:** *Question:* Do associations between depressive symptom trajectories and incident cognitive impairment vary by the rigor of outcome ascertainment?

*Findings:* In this cohort study of 91,043 adults from 4 international cohorts, effect sizes increased stepwise across a 5-level outcome ascertainment hierarchy. Only incident-worsening trajectories were detectable under nonadjudicated outcomes, while clinical consensus diagnosis revealed independent associations for all non-low trajectories (ORs, 1.66-2.29).

*Meaning:* Prevention trials and burden projections that rely on self-reported outcomes may underestimate the potential benefit of treating depression.

## Introduction

Dementia affects more than 55 million people worldwide, with prevalence projected to triple by 2050 as populations age.^1,2^ No disease-modifying therapy has reduced dementia incidence at the population level, underscoring the urgency of identifying modifiable risk factors with established intervention pathways.^3,4^

Major depressive disorder is one such factor. Meta-analyses estimate pooled relative risks of 1.5 to 2.0 for dementia, attenuating when early follow-up years susceptible to reverse causation are excluded.^5,6^ Plausible mechanisms include vascular and inflammatory pathways, hypothalamic-pituitary-adrenal axis dysregulation, and behavioral correlates such as physical inactivity,^7,8^ and because depression is treatable with interventions of demonstrated effectiveness, it is a rare point of leverage in dementia prevention.^9,10^ Yet global burden and prevention frameworks apply pooled depression-dementia relative risks across populations regardless of how each study ascertained dementia.^4,11^

Longitudinal studies have characterized temporal patterns of depressive symptoms in relation to cognitive outcomes.^12,13^ Hou et al^14^ synthesized exposure across the same four cohorts as cumulative burden, duration, and variability, finding that worsening trajectories predicted dementia; however, that analysis used self-reported outcomes without clinical adjudication and a single outcome definition per cohort. Whether the prognostic value of specific symptom dynamics—recent worsening vs sustained elevation—depends on how outcomes are ascertained has not been tested.

We therefore conducted a harmonized analysis of four international cohorts^15–18^ to examine depressive symptom trajectories in relation to incident cognitive impairment, re-estimating associations across a 5-level outcome ascertainment hierarchy. We hypothesized that (1) incident worsening would be detectable under outcome definitions incorporating cognitive testing, whereas sustained elevation would emerge primarily under clinical adjudication; and (2) effect estimates would increase systematically with ascertainment rigor, explaining between-cohort heterogeneity.

## Methods

### Study Design and Population

This harmonized cohort study used four nationally representative longitudinal aging cohorts: the China Health and Retirement Longitudinal Study (CHARLS), the Health and Retirement Study (HRS), the English Longitudinal Study of Ageing (ELSA), and the Survey of Health, Ageing and Retirement in Europe (SHARE). Baseline (anchor) waves were 2011 (CHARLS), 2010 (HRS), 2006 (ELSA), and 2010 (SHARE). Inclusion criteria were age 50 years or older at baseline, at least one depressive symptom assessment during the exposure window, and absence of cognitive impairment or dementia at baseline. The analytic sample comprised 91,043 participants (7,719 CHARLS; 18,215 HRS; 9,016 ELSA; 56,093 SHARE) (eFigure 1 in the Supplement). All cohorts received ethical approval and obtained written informed consent; this secondary analysis of anonymized public data was approved by the Ethics Committee of Tongde Hospital of Zhejiang Province (2025-046[K]) and followed the Declaration of Helsinki.

### Temporal Architecture

Trajectory classes were derived from the full exposure window in each cohort (CHARLS, 2011-2015; HRS, 2000-2022; ELSA, 2006-2018; SHARE, 2004-2021), and follow-up began at the landmark wave (CHARLS, 2015; HRS, 2010; ELSA, 2018; SHARE, 2010). For HRS and SHARE the exposure window extends beyond the landmark, so classification incorporates post-landmark symptom information. Two internal checks argue against systematic information leakage or reverse causation: the persistent-high trajectory, defined by the same post-landmark information, was null under soft outcomes in both cohorts, and lagged findings in CHARLS were consistent with primary findings (HR, 1.38 vs 1.31). The full argument, including why pre-anchor sensitivity analyses were not feasible, is detailed in the eMethods in the Supplement.

### Depressive Symptom Assessment

Depressive symptoms were assessed with the 10-item Center for Epidemiologic Studies Depression Scale (CES-D 10) in CHARLS,^22^ the 8-item CES-D in HRS and ELSA,^23^ and the 12-item EURO-D in SHARE.^24^ Total scores were computed following validated algorithms (range, 0-30, 0-8, and 0-12, respectively); higher scores indicate greater symptom burden. Cross-scale measurement invariance was assessed using multi-group confirmatory factor analysis with alignment optimization (Mplus) to support comparability of trajectory classes across cohorts.^26^

### Trajectory Modeling

Group-based trajectory modeling was used to identify trajectory classes over the exposure window.^19–21^ Models were fitted using the hlme function in the lcmm R package with linear time terms, random intercepts, and 1 to 4 latent classes.^25^ Model selection used the Bayesian Information Criterion (BIC), average posterior probability (>= 0.70), and minimum class size (>= 2%); class identity was verified using shape mean tables. Labels (persistent low, persistent high, incident worsening, remitting) were assigned by trajectory shape. Participants were assigned to their most likely class (modal assignment); posterior probability weighting yielded similar results. Model selection details are in the eMethods.

### Cognitive Outcomes

The primary outcome was incident cognitive impairment, operationalized hierarchically by outcome ascertainment rigor: level 1 (self-reported dementia combined with cognitive decline: HRS soft outcome = self-reported dementia or >= 3-point decline in the 27-point cognitive composite; ELSA = sustained >= 1 SD decline in the cognitive composite across >= 2 consecutive waves, referenced to baseline distribution); level 2 (composite of cognitive testing and self-report: CHARLS = >= 4-point decline in the 28-point cognitive total score or incident self-reported memory-related disease); level 3 (dementia diagnosis or cognitive decline: SHARE = first-recorded dementia diagnosis or >= 1 SD decline in the 24-point cognitive composite); level 4 (screening instruments: SHARE-HCAP = Jorm IQCODE >= 3.3 or CSID informant score >= 10, assessed at wave 9 [2021]); and level 5 (clinical consensus diagnosis: HRS-HCAP = vs1hcapdx algorithm incorporating neuropsychological testing, informant interview, and physician adjudication, assessed at the 2016 wave).^27,28^ The HRS-HCAP outcome was available in a subsample (n = 3,196); SHARE-HCAP in n = 1,405. Memory change was assessed as continuous z-score change from 2016 to 2022 in HRS-HCAP participants with repeated testing (n = 1,492).

### Covariates

Covariates assessed at baseline included age, sex, education, marital status, smoking, alcohol consumption, hypertension, diabetes, stroke, heart disease, arthritis, and psychiatric conditions. Models were adjusted in three tiers (demographics; plus education, marital status, smoking, and alcohol; plus comorbidities). Psychiatric conditions were included for CHARLS and HRS only; exclusion yielded similar estimates. Covariate availability varied by cohort (eTable 2 in the Supplement).

### Statistical Analysis

Cox proportional hazards models with age as the time scale estimated associations between trajectory class and incident cognitive impairment, with delayed entry at the landmark wave; proportional hazards assumptions were verified using Schoenfeld residuals.^34^ The primary analysis was unweighted; survey-weighted CHARLS estimates are reported as sensitivity analyses, and the SHARE weighting rationale is in the eMethods. Competing risks from death were addressed using Fine-Gray models in HRS.^33^ Linear mixed models estimated trajectory-specific cognitive slopes. Pooled estimates were obtained using random-effects meta-analysis (DerSimonian-Laird);^31^ meta-regression with ascertainment rigor (levels 1-5) as a continuous moderator tested heterogeneity explanations. Odds ratios from HCAP logistic models approximate hazard ratios given moderate event proportions; robust Poisson regression yielded similar estimates. Klemera-Doubal method (KDM)^29,30^ biological age acceleration was computed using two algorithms, with no trajectory differences observed (methods and results in the eMethods). Missing covariate data were handled by complete-case analysis; trajectory modeling used all available symptom data under the missing-at-random assumption.No formal sample size calculation was performed; all eligible participants were included, and with 903 to 6,846 outcome events per cohort, the study had greater than 90% power to detect a hazard ratio of 1.30 or greater for the incident-worsening trajectory. Analyses were performed in R version 4.3.3 (lcmm, survival, and metafor packages).^25,32^ Two-sided P < .05 was considered significant.

### Sensitivity Analyses

Prespecified sensitivity analyses included lagged models excluding events within 4 years of baseline (CHARLS), survey weighting (CHARLS), a 4-point trajectory including the 2020 wave (CHARLS), competing-risk models (HRS), continuous cognitive slopes, dementia-only HRS-HCAP outcomes, alternative KDM specifications, posterior probability weighting, and covariate exclusions (eTable 1 in the Supplement). A completed STROBE checklist is provided in the Supplement.

## Results

### Cohort Characteristics and Follow-up

The analytic sample comprised 7,719 CHARLS, 18,215 HRS, 9,016 ELSA, and 56,093 SHARE participants, free of cognitive impairment or dementia at baseline (Table 1). Median follow-up from the landmark wave was 5 years (CHARLS and ELSA), 9 years (SHARE), and 10 years (HRS). Incident cognitive impairment or dementia events ranged from 903 (CHARLS) to 6,846 (SHARE).

**Table 1.** Baseline Characteristics by Cohort.

| Characteristic | CHARLS (n = 7,719) | HRS (n = 18,215) | ELSA (n = 9,016) | SHARE (n = 56,093) |
|---|---|---|---|---|
| Age, mean (SD), y | 59.2 (9.8) | 66.8 (10.2) | 66.3 (10.1) | 64.5 (10.3) |
| Female, No. (%) | 4,085 (52.9) | 10,742 (59.0) | 4,861 (53.9) | 32,847 (58.6) |
| Education, mean (SD), y | 5.8 (4.2) | 12.9 (3.1) | 11.2 (2.8) | 10.8 (4.5) |
| Married/partnered, No. (%) | 6,523 (84.5) | 12,847 (70.5) | 6,324 (70.1) | 38,156 (68.0) |
| Current smoker, No. (%) | 2,156 (27.9) | 2,368 (13.0) | 1,113 (12.3) | 10,247 (18.3) |
| Alcohol consumption, No. (%) | 2,945 (38.2) | 10,847 (59.6) | 6,847 (76.0) | 10,268 (18.4) |
| Hypertension, No. (%) | 3,247 (42.1) | 9,847 (54.1) | 3,083 (34.2) | 28,047 (50.0) |
| Diabetes, No. (%) | 1,156 (15.0) | 3,247 (17.8) | 657 (7.3) | 8,414 (15.0) |
| Stroke, No. (%) | 463 (6.0) | 1,395 (7.7) | 541 (6.0) | 2,805 (5.0) |
| Heart disease, No. (%) | 1,027 (13.4) | 3,651 (20.0) | 1,803 (20.0) | 9,537 (17.0) |
| Arthritis, No. (%) | 2,639 (34.2) | 8,651 (47.5) | — | 13,701 (24.4) |
| Psychiatric conditions, No. (%) | 386 (5.0) | 3,100 (17.0) | — | — |
| Baseline CES-D/EURO-D, mean (SD) | 3.2 (3.1) | 2.1 (2.4) | 2.4 (2.6) | 2.8 (2.9) |
| Follow-up from landmark, median (IQR), y | 5 (4-5) | 10 (8-12) | 5 (4-5) | 9 (7-11) |
| Incident cognitive impairment/dementia events, No. | 903 | 2,793 | 1,999 | 6,846 |

**Note:** Values are presented as mean (SD) or No. (%) unless otherwise indicated. ELSA lacked data on arthritis and psychiatric conditions; SHARE lacked data on psychiatric conditions. Percentages for variables with missing responses were computed on nonmissing denominators. Current smoking was defined as any current smoking (daily cigarettes > 0); diabetes as ever physician-diagnosed diabetes.

### Depressive Symptom Trajectories

Group-based trajectory modeling identified distinct longitudinal patterns in all four cohorts (Figure 1). In CHARLS (3 waves), a 3-class solution was optimal (BIC, 84,427): persistent low (75.9%), persistent high (15.4%), and incident worsening (8.7%). In HRS (12 waves), ELSA (7 waves), and SHARE (8 waves), 4-class solutions were consistently supported (BIC, 546,376, 156,476, and 857,923, respectively), each comprising persistent-low (72.0%-77.6%), persistent-high (4.4%-5.4%), incident-worsening (8.4%-11.9%), and remitting (9.2%-10.7%) classes. The incident-worsening class showed low baseline symptoms followed by sustained escalation.

**Figure 1.**
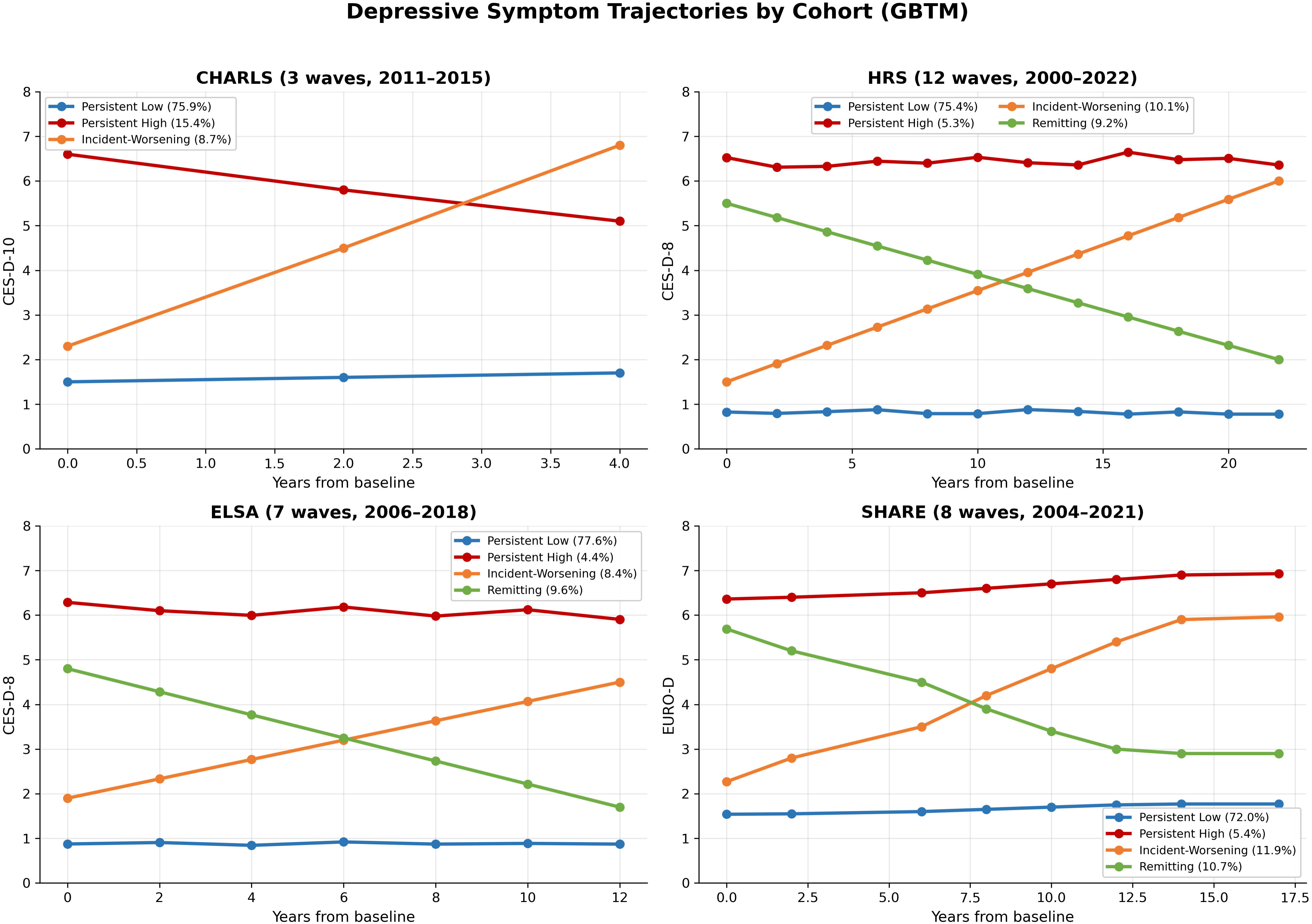
Group-based trajectory modeling identified distinct longitudinal patterns of depressive symptoms in each cohort. CHARLS (3 waves, 2011-2015): 3-class solution (persistent low, 75.9%; persistent high, 15.4%; incident worsening, 8.7%). HRS (12 waves, 2000-2022), ELSA (7 waves, 2006-2018), and SHARE (8 waves, 2004-2021): 4-class solutions with persistent low, persistent high, incident worsening, and remitting classes. The incident-worsening class is characterized by low baseline symptoms followed by sustained escalation.

### Trajectories and Incident Cognitive Impairment

In Cox models adjusted for demographics, health behaviors, and comorbidities, the incident-worsening trajectory was significantly associated with incident cognitive impairment in CHARLS (HR, 1.31; 95% CI, 1.06-1.62) and SHARE (HR, 1.99; 95% CI, 1.87-2.13) but not HRS (HR, 1.05; 95% CI, 0.95-1.16) or ELSA (HR, 1.10; 95% CI, 0.95-1.27) (Table 2). The persistent-high trajectory was not significantly associated in any cohort using soft outcomes (HR range, 1.07-1.15), and the remitting trajectory was null (HR, 1.00-1.13).

**Table 2.** Trajectory Classes and Incident Cognitive Impairment.

| Cohort | Trajectory class | No. (%) | Model 1 HR (95% CI) | Model 2 HR (95% CI) | Model 3 HR (95% CI) | P value |
|---|---|---|---|---|---|---|
| CHARLS | Persistent low | 5,856 (75.9) | Reference | Reference | Reference | — |
|  | Persistent high | 1,192 (15.4) | 1.15 (0.94-1.41) | 1.12 (0.91-1.37) | 1.11 (0.91-1.36) | .29 |
|  | Incident worsening | 671 (8.7) | 1.35 (1.09-1.67) | 1.32 (1.07-1.64) | 1.31 (1.06-1.62) | .01 |
| HRS | Persistent low | 13,734 (75.4) | Reference | Reference | Reference | — |
|  | Persistent high | 965 (5.3) | 1.08 (0.95-1.23) | 1.07 (0.94-1.22) | 1.07 (0.94-1.22) | .31 |
|  | Incident worsening | 1,840 (10.1) | 1.06 (0.96-1.17) | 1.05 (0.95-1.16) | 1.05 (0.95-1.16) | .35 |
|  | Remitting | 1,676 (9.2) | 1.02 (0.92-1.13) | 1.01 (0.91-1.12) | 1.00 (0.90-1.11) | .92 |
| ELSA | Persistent low | 7,000 (77.6) | Reference | Reference | Reference | — |
|  | Persistent high | 397 (4.4) | 1.18 (0.98-1.42) | 1.16 (0.96-1.40) | 1.15 (0.95-1.39) | .16 |
|  | Incident worsening | 757 (8.4) | 1.12 (0.97-1.29) | 1.11 (0.96-1.28) | 1.10 (0.95-1.27) | .22 |
|  | Remitting | 862 (9.6) | 1.14 (0.99-1.31) | 1.13 (0.98-1.30) | 1.13 (0.98-1.30) | .09 |
| SHARE | Persistent low | 40,387 (72.0) | Reference | Reference | Reference | — |
|  | Persistent high | 3,029 (5.4) | 1.08 (1.01-1.15) | 1.07 (1.00-1.14) | 1.07 (1.00-1.14) | .05 |
|  | Incident worsening | 6,675 (11.9) | 2.01 (1.89-2.14) | 2.00 (1.88-2.13) | 1.99 (1.87-2.13) | <.001 |
|  | Remitting | 6,002 (10.7) | 1.01 (0.95-1.08) | 1.00 (0.94-1.07) | 1.00 (0.94-1.07) | .89 |

**Note:** Model 1: adjusted for age and sex. Model 2: additionally adjusted for education, marital status, smoking, and alcohol. Model 3: additionally adjusted for comorbidities (hypertension, diabetes, stroke, heart disease, arthritis, and psychiatric conditions where available). HR indicates hazard ratio.

### Outcome Ascertainment Gradient and HCAP Analyses

Effect sizes for the incident-worsening trajectory increased stepwise with ascertainment rigor (Figure 2): weakest for self-report plus decline (level 1: HRS HR, 1.05; ELSA HR, 1.10), stronger for composite outcomes (level 2: CHARLS HR, 1.31; level 3: SHARE HR, 1.99), stronger still for screening instruments (level 4: SHARE-HCAP OR, 2.37; 95% CI, 1.34-4.20), and significant for clinical consensus diagnosis (level 5: HRS-HCAP OR, 1.71; 95% CI, 1.33-2.20). The level 4-5 inversion was within confidence interval overlap and reflected the small, cross-sectional SHARE-HCAP subsample (n = 1,405).

**Figure 2.**
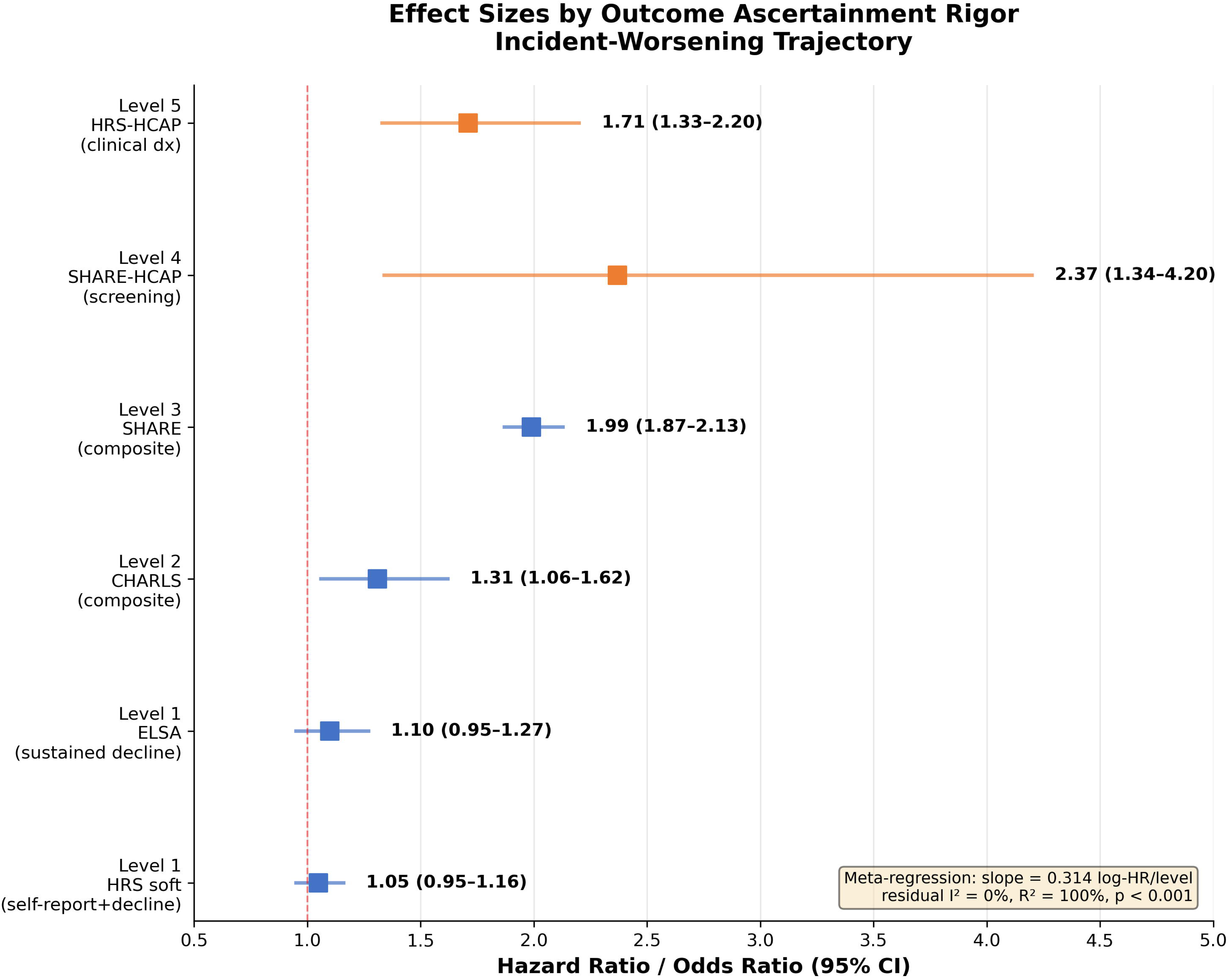
Forest plot showing hazard ratios (HR) and odds ratios (OR) for the association between the incident-worsening depressive symptom trajectory and incident cognitive impairment, stratified by outcome ascertainment level (level 1 = self-report; level 2 = composite with cognitive testing; level 3 = dementia diagnosis or decline; level 4 = screening instruments; level 5 = clinical consensus diagnosis). Effect sizes increase stepwise with ascertainment rigor. Meta-regression slope: 0.314 log HR per level (P < .001), with negligible residual heterogeneity (I^2^ = 0%, R^2^ = 100%).

Under clinical adjudication (HRS-HCAP; n = 3,196), all non-low trajectories were independently associated with mild cognitive impairment or dementia: incident worsening (OR, 1.71; 95% CI, 1.33-2.20), persistent high (OR, 2.29; 95% CI, 1.51-3.46), and remitting (OR, 1.66; 95% CI, 1.25-2.21). For adjudicated dementia, ORs were 2.10 (95% CI, 1.46-3.01), 3.32 (95% CI, 1.85-5.95), and 1.69 (95% CI, 1.06-2.69), respectively. Memory change scores from 2016 to 2022 (n = 1,492) declined by approximately 0.6 SD across all trajectory groups with no between-group differences.

### Pooled Analysis

Random-effects meta-analysis yielded a pooled HR of 1.32 (95% CI, 0.90-1.94) with substantial heterogeneity (I^2^ = 97.8%) (Table 3). Substituting the HRS-HCAP adjudicated outcome increased the pooled estimate to 1.49 (95% CI, 1.05-2.12; I^2^ = 95.1%). Meta-regression with ascertainment rigor as moderator left negligible residual heterogeneity (slope, 0.314 log HR per level; P < .001; residual I^2^ = 0%; R^2^ = 100%), although this analysis was exploratory (4 cohorts).

**Table 3.**
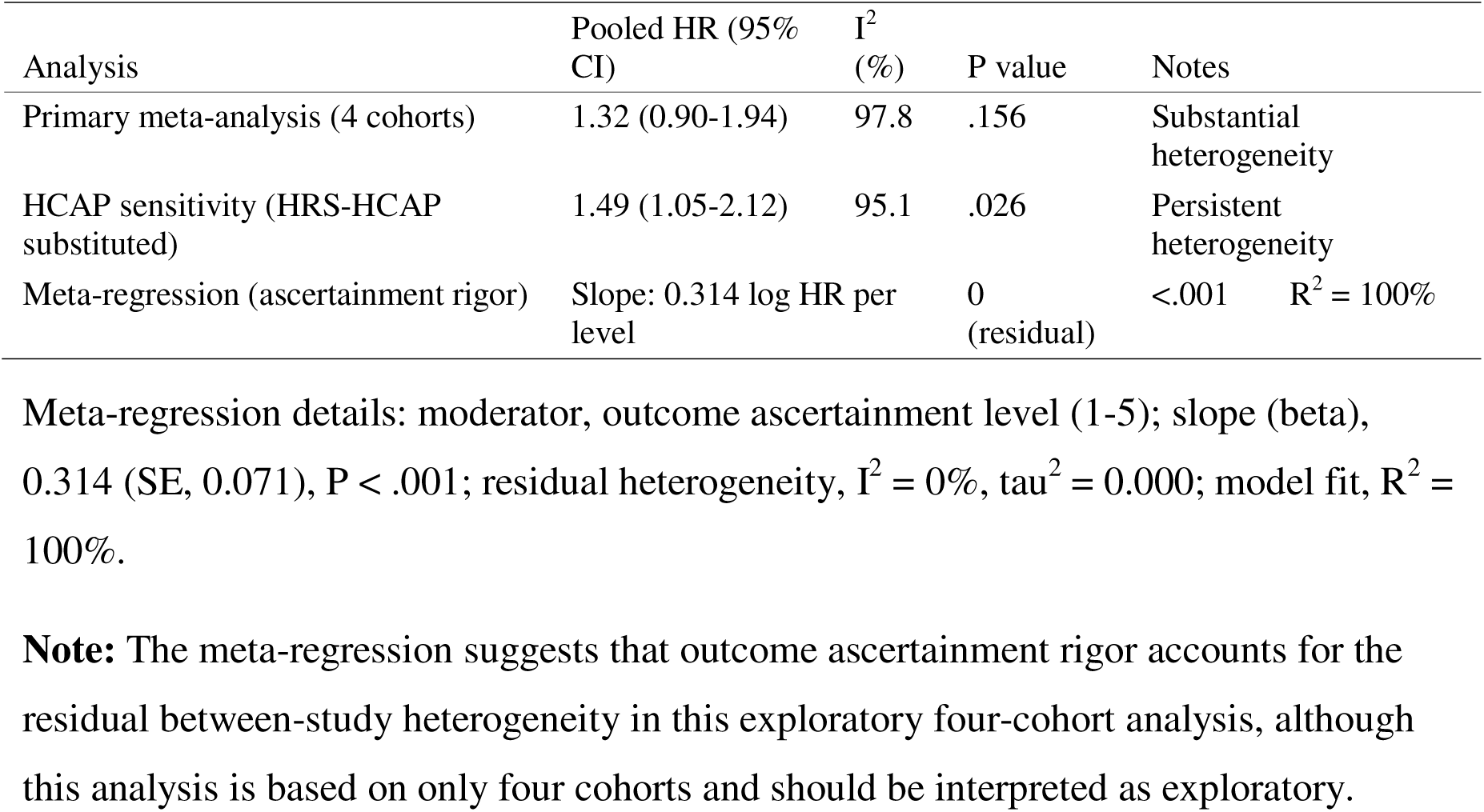
Pooled Meta-Analysis and Meta-Regression.

| Analysis | Pooled HR (95% CI) | I<sup>2</sup> (%) | P value | Notes |
|---|---|---|---|---|
| Primary meta-analysis (4 cohorts) | 1.32 (0.90-1.94) | 97.8 | .156 | Substantial heterogeneity |
| HCAP sensitivity (HRS-HCAP substituted) | 1.49 (1.05-2.12) | 95.1 | .026 | Persistent heterogeneity |
| Meta-regression (ascertainment rigor) | Slope: 0.314 log HR per level |  | 0 (residual) | <.001 R<sup>2</sup> = 100% |

Meta-regression details: moderator, outcome ascertainment level (1-5); slope (beta), 0.314 (SE, 0.071), P < .001; residual heterogeneity, I<sup>2</sup> = 0%, tau<sup>2</sup> = 0.000; model fit, R<sup>2</sup> = 100%.
**Note:** The meta-regression suggests that outcome ascertainment rigor accounts for the residual between-study heterogeneity in this exploratory four-cohort analysis, although this analysis is based on only four cohorts and should be interpreted as exploratory.

### Biological Aging and Sensitivity Analyses

KDM biological age acceleration did not differ across trajectories in CHARLS (n = 4,814; eMethods in the Supplement). Findings were robust to lagged analysis (CHARLS HR, 1.38; 95% CI, 1.06-1.78), survey weighting (HR, 1.37; 95% CI, 1.07-1.77), the 2020-wave trajectory (HR, 1.17; 95% CI, 0.94-1.46), and competing-risk models (eTable 1). Linear mixed models showed significant incident-worsening-by-slope interactions in CHARLS (−0.144 points/year; 95% CI, −0.211 to −0.077) and ELSA (−0.076 points/2 years; 95% CI, −0.094 to −0.057) despite null Cox results in ELSA.

## Discussion

In this harmonized analysis of 91,043 older adults from four international cohorts, the association between depressive symptom trajectories and incident cognitive impairment depended systematically on how outcomes were ascertained. Under nonadjudicated outcomes, only recent symptom worsening was detectable, whereas clinical consensus diagnosis revealed independent risk for all non-low trajectories, with the largest estimate for persistent elevation predicting adjudicated dementia (OR, 3.32). Ascertainment rigor statistically explained all residual between-cohort heterogeneity in an exploratory meta-regression. The stepwise gradient—from self-report (HR, 1.05-1.10) to composite outcomes (HR, 1.31-1.99) to screening instruments (OR, 2.37) to consensus diagnosis (OR, 1.71)—suggests that capacity to detect depression-related cognitive risk varies with outcome measurement: self-reported outcomes may misclassify mild impairment as intact, while adjudication incorporating neuropsychological testing and informant reports captures impairment that self-report misses.^27,28^ The apparent specificity of incident worsening in soft-outcome analyses may therefore reflect limited sensitivity of these measures for sustained or remitting patterns rather than the biological irrelevance of those patterns.

Our findings extend the work of Hou et al,^14^ who found that worsening depression exposure predicted dementia in the same four cohorts using self-reported outcomes but did not test measurement dependence. That we reproduce their core finding while showing it to be measurement-contingent reconciles seemingly discordant literature: studies using self-report or composite outcomes would detect only the most dramatic symptom dynamics, whereas adjudicated studies would detect subtler patterns. This is consistent with earlier trajectory studies, including the “high and increasing” trajectory identified by Kaup et al^12^ and a recent 23-year UK cohort linking specific midlife depressive symptoms to dementia.^36^ Reverse-causation and leakage considerations are detailed in the Methods and eMethods.

The dissociation between significant cognitive-slope interactions (CHARLS, ELSA) and null event-based associations in ELSA suggests that continuous outcomes capture gradual decline that threshold-based event definitions miss. Null memory-change findings in HRS-HCAP may reflect selective survival of slower decliners into the 2022 assessment wave. The absence of trajectory differences in KDM biological aging acceleration in CHARLS suggests the incident-worsening association is not accounted for by cumulative systemic aging, although other biological pathways cannot be excluded.^7,8^

These findings have implications for dementia prevention. If depression-related cognitive risk is underestimated under soft outcomes, prevention trials and burden projections built on such outcomes may systematically understate the benefit of depression treatment, identified by the 2024 Lancet Commission as a prevention strategy.^4^ The fraction of dementia attributable to depression—and hence the potential population impact of scaling up treatment—may be larger than estimated under self-report-based outcomes.^9,10^ We did not model treatment coverage directly; quantifying averted burden under scaled treatment requires a treatment-adjusted attributable fraction framework, which we did not attempt.

Strengths include the harmonized four-cohort design spanning diverse health systems, the prespecified ascertainment hierarchy, clinically adjudicated outcomes, and systematic heterogeneity testing. Limitations include the small number of cohorts for meta-regression (k = 4), rendering the R^2^ = 100% finding exploratory; the cross-sectional SHARE-HCAP analysis; mixing of odds ratios and hazard ratios across levels; outcome operationalization heterogeneity; classification of ELSA’s sustained-decline outcome as level 1 despite incorporating repeated cognitive testing; restriction of KDM analyses to CHARLS; potential information leakage from full-window trajectory classification in HRS and SHARE, which cannot be fully excluded; and absence of treatment coverage data.

Depressive symptom trajectories are associated with incident cognitive impairment, but detectability and magnitude depend on outcome ascertainment. Recent worsening was the only pattern detectable under less rigorous definitions, while sustained elevation and remitting patterns emerged under clinical adjudication. Outcome measurement may explain substantial heterogeneity across studies of depression and dementia risk; trials and prevention strategies should incorporate rigorous cognitive outcome assessment to avoid underestimating the potential benefit of depression treatment.

## Supporting information

eSupplement

## Data Availability

Individual-level deidentified analytic data and statistical code will be available from the corresponding author upon publication, for any purpose, without investigator support, subject to a signed data access agreement. Source cohort data (CHARLS, HRS, ELSA, and SHARE) are available to bona fide researchers through each study's own data access procedures (charls.pku.edu.cn; hrs.isr.umich.edu; elsa-project.ac.uk; share-eric.eu).

## Author Contributions

Drs Liu and Guo had full access to all of the data in the study and take responsibility for the integrity of the data and the accuracy of the data analysis. Dr Liu: conceptualization, data analysis, visualization, writing—original draft. Dr Guo: conceptualization, methodology, supervision, writing—review and editing. Both authors directly accessed and verified the underlying data at the individual-participant level and accept responsibility to submit for publication.

## Additional Contributions

During the preparation of this work, the authors used Kimi (Moonshot AI) to assist with drafting the cover letter and language editing. The authors reviewed, verified, and edited all content and take full responsibility for the integrity and accuracy of the publication.

