## Supplementary material for "Depressive Symptom Trajectories and Incident Cognitive Impairment by Outcome Ascertainment": eSupplement

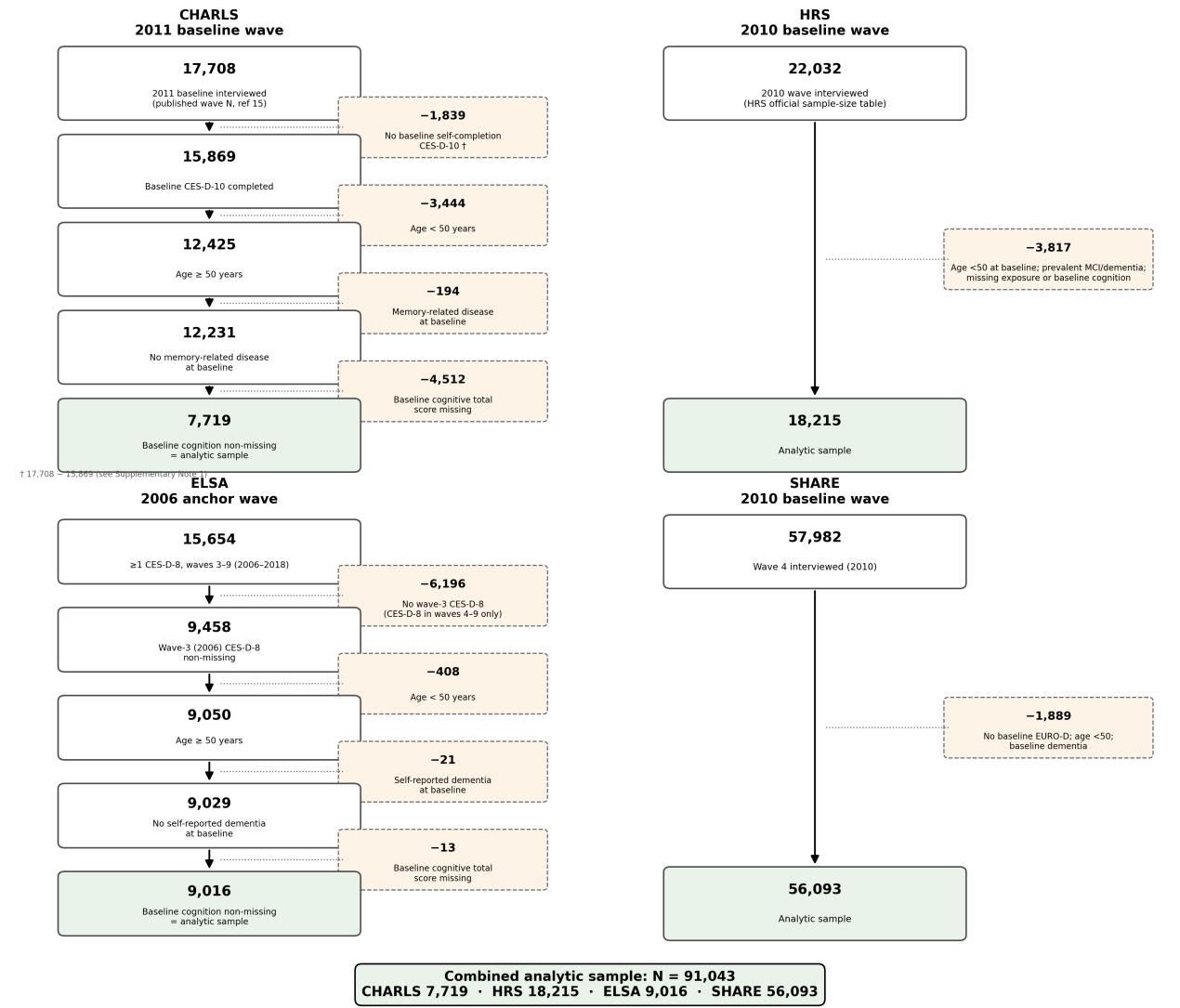

### eFigure 1. Study Flow Diagram. CHARLS: of 17,708 participants interviewed at the 2011 baseline wave, 1,839 did not complete the baseline self-completion CES-D-10, 3,444 were aged < 50 years, 194 had prevalent memory-related disease at baseline, and 4,512 lacked a baseline cognitive total score, leaving 7,719. HRS: of 22,032 participants interviewed in 2010 (per the HRS official sample-size table), 3,817 were excluded (age < 50 years at baseline, prevalent MCI or dementia, or missing exposure or baseline cognition), leaving 18,215. ELSA: 15,654 participants had at least 1 CES-D-8 observation during waves 3-9 (2006-2018); 6,196 did not complete the wave-3 (2006) CES-D-8, 408 were aged < 50 years, 21 reported dementia at baseline, and 13 lacked a baseline cognitive score, leaving 9,016. SHARE: of 57,982 participants interviewed at wave 4 (2010), 1,889 lacked baseline EURO-D, were aged < 50 years, or had baseline dementia, leaving 56,093. Combined analytic sample: N = 91,043.

### eMethods

#### eMethods 1. Temporal Architecture and Information-Leakage Considerations

Trajectory classes were derived from the full exposure window in each cohort (CHARLS: 2011-2015; HRS: 2000-2022; ELSA: 2006-2018; SHARE: 2004-2021). Participants entered the risk period at the landmark wave (CHARLS: 2015; HRS: 2010; ELSA: 2018; SHARE: 2010). For HRS and SHARE, the exposure window extends beyond the landmark; trajectory classification therefore incorporates post-landmark symptom information. Sensitivity analyses restricting to pre-landmark waves were not feasible: SHARE pre-anchor waves supported only a 2-class solution, and HRS pre-anchor waves (2000-2008) supported only a 3-class solution lacking a persistent-high class. If information leakage systematically inflated associations, the persistent-high trajectory—defined by the same post-landmark information—should also show elevated risk in HRS and SHARE; however, persistent-high associations were null in both cohorts under soft outcomes. The consistency of lagged findings in CHARLS (HR, 1.38 vs 1.31 in the primary analysis) argues against reverse causation as the primary explanation, although this strongest test could not be replicated in the leakage-affected cohorts because pre-landmark waves were insufficient for trajectory classification. For HRS-HCAP analyses, the adjudicated outcome was assessed at the 2016 wave, overlapping the later exposure window.

#### eMethods 2. Survey Weighting

The primary analysis was unweighted. Survey-weighted estimates for CHARLS (wtresp) are reported in eTable 1. HRS and ELSA were not weighted because their sampling frames have near-complete baseline coverage. SHARE longitudinal weights were available for only 3,463 participants (6.2%) who completed all waves from 1 to 9; restricting to this subset would introduce selection bias toward long-term survivors, so SHARE analyses were unweighted.

#### eMethods 3. Trajectory Model Selection Details

Models were fitted using the hlme function in the lcmm R package with linear time terms, random intercepts (growth mixture model specification), and 1 to 4 latent classes. Model selection was based on Bayesian Information Criterion (BIC), average posterior probability (AvePP >= 0.70), and minimum class size (>= 2%). BIC values were 84,427 (CHARLS, 3-class solution), 546,376 (HRS, 4-class), 156,476 (ELSA, 4-class), and 857,923 (SHARE, 4-class). Class identity was verified using shape mean tables (class-specific mean symptom scores at each wave); labels were assigned based on trajectory shape, not class numbering.

#### eMethods 4. KDM Biological Age Methods and Results

KDM biological age acceleration was computed using two algorithms: (1) a formal 6-biomarker KDM (log-CRP, log-creatinine, HbA1c, total cholesterol, blood urea nitrogen, systolic blood pressure), adapted from the KDM-EB algorithm by excluding biomarkers unavailable in CHARLS (FEV1, albumin, alkaline phosphatase); and (2) a simplified 7-biomarker variant (mean corpuscular volume, white blood cell count, hemoglobin, creatinine, CRP, HbA1c, cystatin C), a residual-weighted exploratory version. Acceleration was defined as predicted biological age minus chronological age. KDM biological age acceleration did not differ across depressive symptom trajectories in CHARLS (formal 6-biomarker KDM: mean acceleration, +0.016, -0.063, and -0.020 years for persistent low, persistent high, and incident worsening, respectively; n = 4,814; eTable 1). The simplified 7-biomarker variant showed no adjusted associations. These findings should be interpreted as specific to the available biomarker set and to the CHARLS population, and do not exclude other biological pathways.

### eTable 1. Sensitivity Analyses

| Sensitivity analysis | Cohort | Effect estimate (95% CI) | P value | Interpretation |
| --- | --- | --- | --- | --- |
| Lagged analysis (4-year exclusion) | CHARLS | 1.38 (1.06-1.78) | .018 | Robust to reverse causation |
| Survey weighting | CHARLS | 1.37 (1.07-1.77) | .014 | Robust to sampling design |
| 4-point trajectory (2020 wave) | CHARLS | 1.17 (0.94-1.46) | .168 | Attenuated but direction consistent |
| Competing-risk model (Fine-Gray) | HRS | sHR 1.04-1.08 | >.05 | Null robust to death competing risk |
| Dementia-only outcome | HRS-HCAP | 2.10 (1.46-3.01) | <.001 | Stronger for dementia vs MCI |
| Posterior probability weighting | All | Similar to modal assignment | — | Robust to class assignment method |
| Excluding psychiatric conditions | CHARLS, HRS | Similar to primary | — | Robust to covariate specification |
| LMM cognitive slope | CHARLS | -0.144 points/year (-0.211 to -0.077) | <.001 | Significant slope difference |
| LMM cognitive slope | ELSA | -0.076 points/2 years (-0.094 to -0.057) | <.001 | Significant despite null Cox |
| Memory change (2016-2022) | HRS-HCAP | Approximately 0 (null) | >.20 | Selective survival explanation |
| KDM biological age acceleration | CHARLS | +0.016, -0.063, -0.020 years | >.05 | No trajectory differences |
| Pre-anchor trajectory classification | HRS | Not feasible (3-class optimal) | — | Lacks persistent-high class |
| Pre-anchor trajectory classification | SHARE | Not feasible (2-class optimal) | — | Insufficient waves for 4-class |

**Note:** sHR indicates subdistribution hazard ratio; LMM, linear mixed model; KDM, Klemera-Doubal method. Effect estimates are for the incident-worsening trajectory (model 3) unless otherwise indicated. The pre-anchor sensitivity analyses were not feasible because the reduced wave count did not support the full 4-class structure (eMethods 1).

### eTable 2. Covariate Availability by Cohort

| Covariate | CHARLS | HRS | ELSA | SHARE |
| --- | --- | --- | --- | --- |
| Age | Yes | Yes | Yes | Yes |
| Sex | Yes | Yes | Yes | Yes |
| Education | Yes | Yes | Yes | Yes |
| Marital status | Yes | Yes | Yes | Yes |
| Smoking | Yes | Yes | Yes | Yes |
| Alcohol consumption | Yes | Yes | Yes | Yes |
| Hypertension | Yes | Yes | Yes | Yes |
| Diabetes | Yes | Yes | Yes | Yes |
| Stroke | Yes | Yes | Yes | Yes |
| Heart disease | Yes | Yes | Yes | Yes |
| Arthritis | Yes | Yes | No | Yes |
| Psychiatric conditions | Yes | Yes | No | No |

**Note:** Psychiatric conditions were included in model 3 for CHARLS and HRS only; sensitivity analyses excluding this covariate yielded similar estimates (eTable 1).

### eAppendix. Sample Construction Accounting

**CHARLS.** The first node (17,708) is the published wave-1 N (“…involved 17,708 respondents” [Zhao et al, *Int J Epidemiol* 2014]). CHARLS wave files are distributed as separate module files; the 2011 Health_Status_and_Functioning module contains 17,596 records, and the analysis wide file links 17,684 participants with at least 1 variable from 2011 across modules. The first exclusion (1,839) is the arithmetic difference between the published wave N and the 15,869 participants completing the baseline self-completion CES-D-10, not a pipeline count. All subsequent exclusions (3,444; 194; 4,512) are pipeline outputs, yielding 7,719.

**HRS.** The first node (22,032) is the total number of 2010 core respondents (Panel 1: 15,749; 2010 new cohort: 6,283; HRS official sample-size documentation). The exclusion (3,817) is the arithmetic difference 22,032 - 18,215.

**ELSA.** The exposure-window sample (15,654) is the number of unique participants with at least one CES-D-8 observation during waves 3-9 (2006-2018) in the harmonized ELSA file. Sample construction occurs at the wave-3 (2006) baseline: 6,196 participants without a wave-3 CES-D-8 (their qualifying observations fall in waves 4-9 only; 6,196 = 15,654 - 9,458, both pipeline outputs), 408 aged less than 50 years, 21 with self-reported dementia, and 13 with missing baseline cognition, yielding 9,016. The full harmonized file contains 21,186 unique participants (waves 1-11), of whom 18,809 have at least 1 CES-D-8 observation in any wave; neither enters the sample construction. No analytic participant entered the study after wave 3.

**SHARE.** The first node (57,982) is the published wave-4 N; the exclusion (1,889) is the arithmetic difference 57,982 - 56,093.
